# IMPACT OF CENTRALIZED NIKSHAY ON TB NOTIFICATION AND DATA COMPLETENESS IN INDIGENOUS TRIBES: A 21-YEAR STUDY

**DOI:** 10.64898/2026.10.13.26365279

**Authors:** Mona Rastogi, Aishwarya S. Babu, R Abhishek, Shruthi Vasanthaiah, Parthasarathy Ponnusamy, N Nagarathna, M Rajesh, Deepak Murthy, Akhilesh Pandey, Kumar Gavali Suryanarayana, Renu Verma

## Abstract

**BACKGROUND:** Indigenous tribes face a disproportionately high tuberculosis (TB) burden due to deep-rooted socioeconomic disadvantage, highlighting the need for strengthened surveillance and control strategies.

**METHODS:** We conducted a 21-year retrospective study (2005-2025) among tribal and neighboring non-tribal populations attending a tribal-serving hospital in Karnataka. Pre-Nikshay data were transcribed from paper registers, and post-Nikshay data were obtained from the digital system. We assessed negative binomial interrupted time-series regression model, data completeness, TB risk factors, and used multivariable logistic regression to identify determinants of poor treatment outcomes and household contact screening yield.

**RESULTS:** We analyzed 1,512 TB cases (tribal = 1,021; non-tribal = 491). TB detection increased significantly over time, with a steeper rise among tribal populations (p<0.001). Nikshay implementation significantly improved data completeness (Wilcoxon W = 9900, p<0.001). Among tribal patients, males had higher odds of poor treatment outcome (aOR 2.81, 95% CI 1.70–4.85; *p*<0.001). Diabetes (aOR 2.10, 95% CI 1.05–4.03; *p*=0.029) was independently associated with poor treatment outcome. Among 503 index cases, 1,535 household contacts were identified and 1,161 (75.6%) screened; coverage was lower among tribal contacts (69.8% vs 87.2%; p<0.001).

**CONCLUSIONS:** Nikshay strengthened TB surveillance and improved assessment of TB burden and risk factors in tribal communities.

## INTRODUCTION

Tuberculosis (TB) remains the leading cause of mortality from a single infectious disease worldwide, with India accounting for approximately 25% of the global burden. An estimated 20-25% of incident TB cases globally remain undiagnosed or unreported, with India contributing approximately 8.8% of this global gap¹. This challenge is particularly pronounced among socially and geographically marginalized populations, including Indigenous tribes, prisoners, and urban slum residents, who face barriers to healthcare and heightened transmission risks² .

Indigenous tribal populations in India, comprising approximately 8.6% of the national population, experience disproportionately high TB incidence, drug resistance, and mortality . A national survey reported pulmonary TB prevalence of 432 per 100,000 among tribal communities, compared with 312 per 100,000 nationally . Multidrug-resistant TB (MDR-TB) prevalence has reached approximately 8.8% among the Saharia tribe in Madhya Pradesh and 12-17% among previously treated tribal patients in Gujarat and Maharashtra. These disparities are associated with structural determinants, including displacement, malnutrition, tobacco and alcohol use, low income, inadequate ventilation, and overcrowded housing .

Karnataka state of India has a Scheduled Tribe population of 4.249 million¹ . Among these communities, the Jenu Kuruba are designated a Particularly Vulnerable Tribal Group (PVTG) by the Ministry of Tribal Affairs¹¹. Other underserved Indigenous communities in the region include the Betta Kuruba (Kadu Kuruba), Yarava, and Soliga¹² ¹ . Conservation-driven displacement from ancestral forest habitats in the Western Ghats to peripheral rural and peri-urban settlements has disrupted traditional livelihoods and socio-environmental systems, potentially altering exposure to infectious diseases, including TB ¹ .

Despite policy recognition of their vulnerability, TB data for Indigenous communities remain limited, fragmented, and non-standardized, restricting accurate assessment of disease burden and risk factors¹ . Effective surveillance requires systematic documentation of occupation, living conditions, substance use, and household contacts. Historically, paper-based hospital registers have suffered from inconsistent completeness and limited stratification. India’s Ni-kshay digital TB reporting system, implemented under the National Tuberculosis Elimination Programme (NTEP), provides opportunities to improve data standardization, accessibility, and completeness.

However, its impact on TB surveillance among Indigenous tribes remains insufficiently evaluated, and tribe-specific burden estimates for Karnataka are scarce.

To address these gaps, we compiled a 21-year dataset (2005-2025) of active TB cases among Indigenous tribal communities attending an NTEP-supported rural secondary-care hospital with a Designated Microscopy Centre (DMC) in Karnataka. Neighboring non-tribal residents served as a comparison group. Records from 2005-2017 were manually extracted from hospital registers, while data from 2018-2025 were retrieved from Ni-kshay. We estimated TB burden across five major Indigenous tribal groups, examined associations with occupation, living conditions, and substance use, and compared case notification and data completeness between paper-based registers and digital surveillance.

## METHODS

### Study setting and rationale

The tribes in this study reside predominantly along the Nagarahole and Bandipur forest ranges of the Western Ghats, India. The Swami Vivekananda Youth Movement (SVYM), a well-known non-governmental organization in Karnataka, provides primary and secondary healthcare to these communities in rural Mysuru through community-based initiatives and the Vivekananda Memorial Hospital (VMH), a 100-bed secondary care hospital. The designated microscopy centre (DMC) at VMH serves nearby tribal and non-tribal communities, enabling comparative assessment of TB burden, comorbidities, nutritional status, and risk factors. SVYM maintains comprehensive longitudinal clinical records, supporting reliable retrospective data collection and follow-up. VMH is also an HIV Comprehensive Care Centre (CCC), established in 2003.

### Ethical statement

Ethical approval was obtained from the Institutional Ethics Committee of the Institute of Bioinformatics (IOB/2nd IEC/2023). Full committee review was exempted, as the study used only retrospective data from SVYM (IERB approval no. VMH_EC/2026-009). All patient identifiers were removed before analysis, and results are reported in aggregate to protect confidentiality.

### Data source

Data came from two sources: (i) paper-based SVYM hospital registers documenting Indigenous tribal and non-tribal patients residing near tribal communities, and (ii) the Nikshay portal, the national web-based TB case management system under India’s National Tuberculosis Elimination Programme (NTEP).

### Study population

All TB cases registered in the SVYM hospital registers between 2005 and 2025 were included, covering both indigenous tribal and non-tribal individuals. Patients transferred to other facilities after registration were retained, with treatment outcomes and follow-up information obtained from the Nikshay portal where available. Diagnosis followed the national TB programme guidelines applicable at the time. Sputum smear microscopy was the primary test from 2005, and CBNAAT was used from 2018. Extrapulmonary TB was diagnosed using clinical, radiological, microbiological, and histopathological findings, as appropriate to the disease site.

### Data from hospital registers (2005-2017)

Clinical records from January 2005 to December 2017 were manually transcribed from paper registers at VMH into a structured electronic database by trained SVYM staff, in collaboration with Institute of Bioinformatics scientists. Socio-demographic and clinical variables were captured for five tribal communities (Jenu Kuruba, Betta Kuruba, Soliga, Yarava, and Paniya), including symptoms, HIV and diabetes comorbidities, sputum smear results, site of disease, case type, treatment category, and outcomes. Entries were cross-validated against original records. Treatment failure, loss to follow-up, and death were classified as unfavorable outcomes.

### Nikshay portal (2018-2025)

For the post-Nikshay period (September 2018-2025), data were extracted from the national portal for patients from the same tribal communities and neighboring non-tribal residents. The portal provided variables analogous to the registers (age, sex, weight, caste, residence, symptoms, comorbidities, TB site, case classification, treatment category, and outcomes). It also captured detailed diagnostic and risk-factor information, including CBNAAT, line probe assays, culture, radiography, addictions, occupation, socio-economic status, household contact history, and contact-tracing data (identified, screened, diagnosed, and initiated on treatment).

### Data cleaning and processing

Data were managed in R version 4.3.2 using tidyverse packages (dplyr 1.1.4, tidyr 1.1.3). Completeness, defined as the proportion of records with valid entries (missing or unknown values counted as incomplete), was compared between hospital records and Nikshay across demographic, clinical, laboratory, and treatment variables. Overall completeness scores were averaged across variables, compared using the Wilcoxon rank-sum test, and visualized with violin plots. Data were stratified by source and year, and TB trends were analyzed by population group, region, demographics, treatment category, and outcomes.

### Statistical analysis

We analyzed data in R version 4.3.2 and Microsoft Excel, with figures generated using ggplot2. All tests were two-sided, and P < 0.05 was considered statistically significant. We compared TB detection across groups, including tribal and non-tribal populations, using negative binomial regression, and assessed the effect of Nikshay implementation using a negative binomial interrupted time-series model. Model fit was evaluated using DHARMa simulated residuals (Kolmogorov-Smirnov, dispersion, and outlier tests).

We then used multivariable logistic regression to examine associations of demographic, clinical, and behavioral factors with TB detection and poor treatment outcome (failure, loss to follow-up, or death; coded 1) versus successful outcome (coded 0). Reference categories were female sex, non-tribal population, HIV-negative status, and non-diabetic status. Results are reported as adjusted odds ratios (aOR) with 95% confidence intervals (CI). Alcohol and other addiction variables were excluded from the primary model because data were available for few patients, and their inclusion would have reduced sample size and increased selection bias. Calibration was assessed using the Hosmer-Lemeshow test. Associations between categorical variables were tested using Pearson’s chi-square test, and continuous measures were compared using the Wilcoxon rank-sum test.

## RESULTS

### Temporal trends in TB detection

A total of 124 tribal hamlets across six regions of Karnataka were included-H.D. Kote (n = 80), Saragur (n = 28), Nanjanagudu/Gundalapete (n = 3), Hunsur/Piriyapatna (n = 11), and Kushalnagara/Somwarpet (n = 2). The majority of hamlets (n = 108) were from H.D. Kote and Saragur combined are directly served by DMC at VMH, Saragur, the nearest secondary care facility for these communities (**Figure 1A**). From hospital registers and Nikshay, we identified 1,512 active TB cases between 2005 and 2025, including 1,021 (67.5%) tribal and 491 (32.5%) non tribal patients. After normalizing case counts to population, TB detection increased significantly over time in both tribal (p = 0.0126) and non tribal (p = 0.0001) groups, but remained consistently higher among tribal communities. (**Figure 1B**)

**Figure 1(A).**
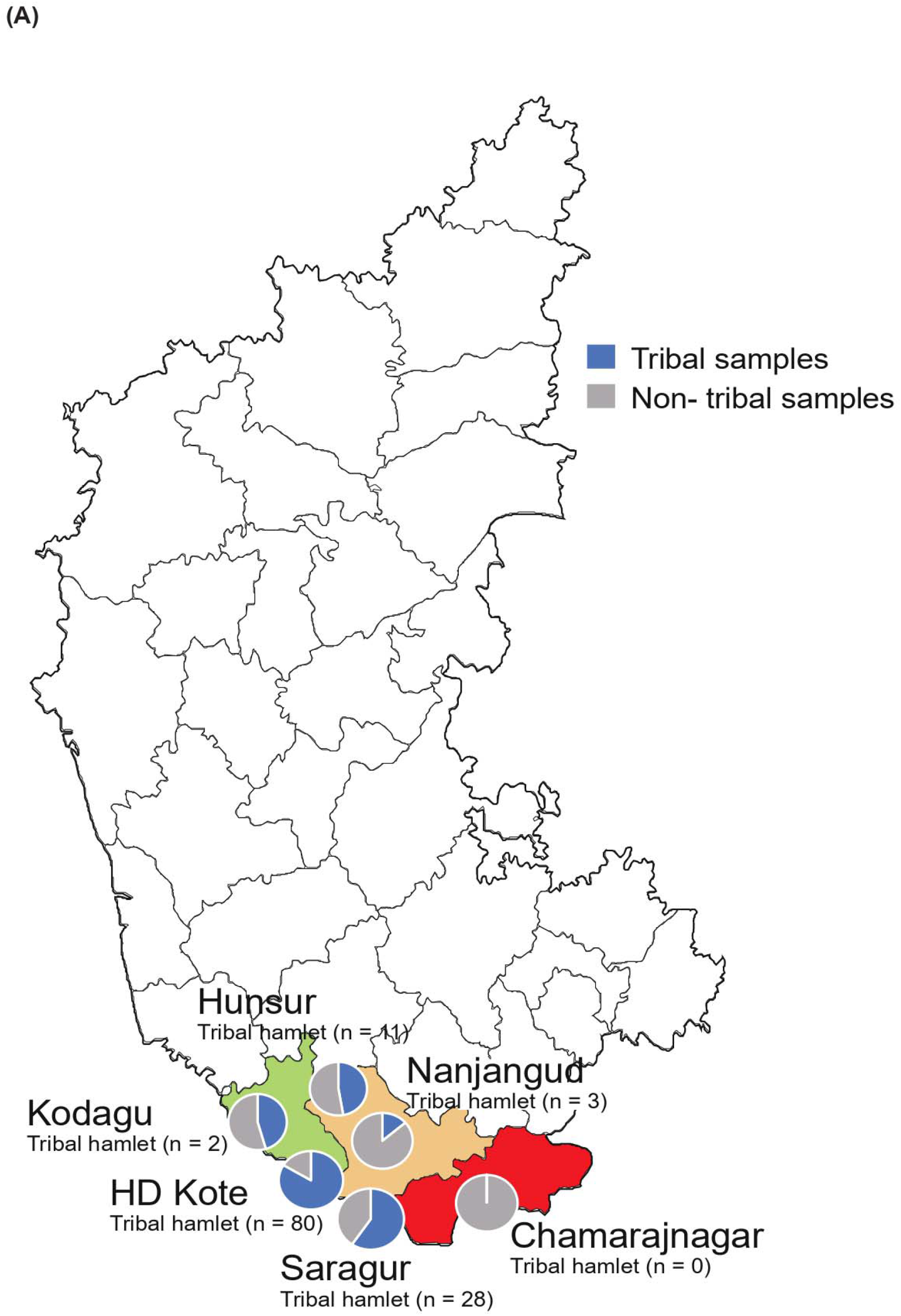
Study area in Karnataka showing the distribution of tribal hamlets and sample collection. The map indicates the number of tribal hamlets within each study zone, while the accompanying pie chart summarizes the numbers of tribal and non-tribal tuberculosis samples included from each zone.

**Figure 1B.**
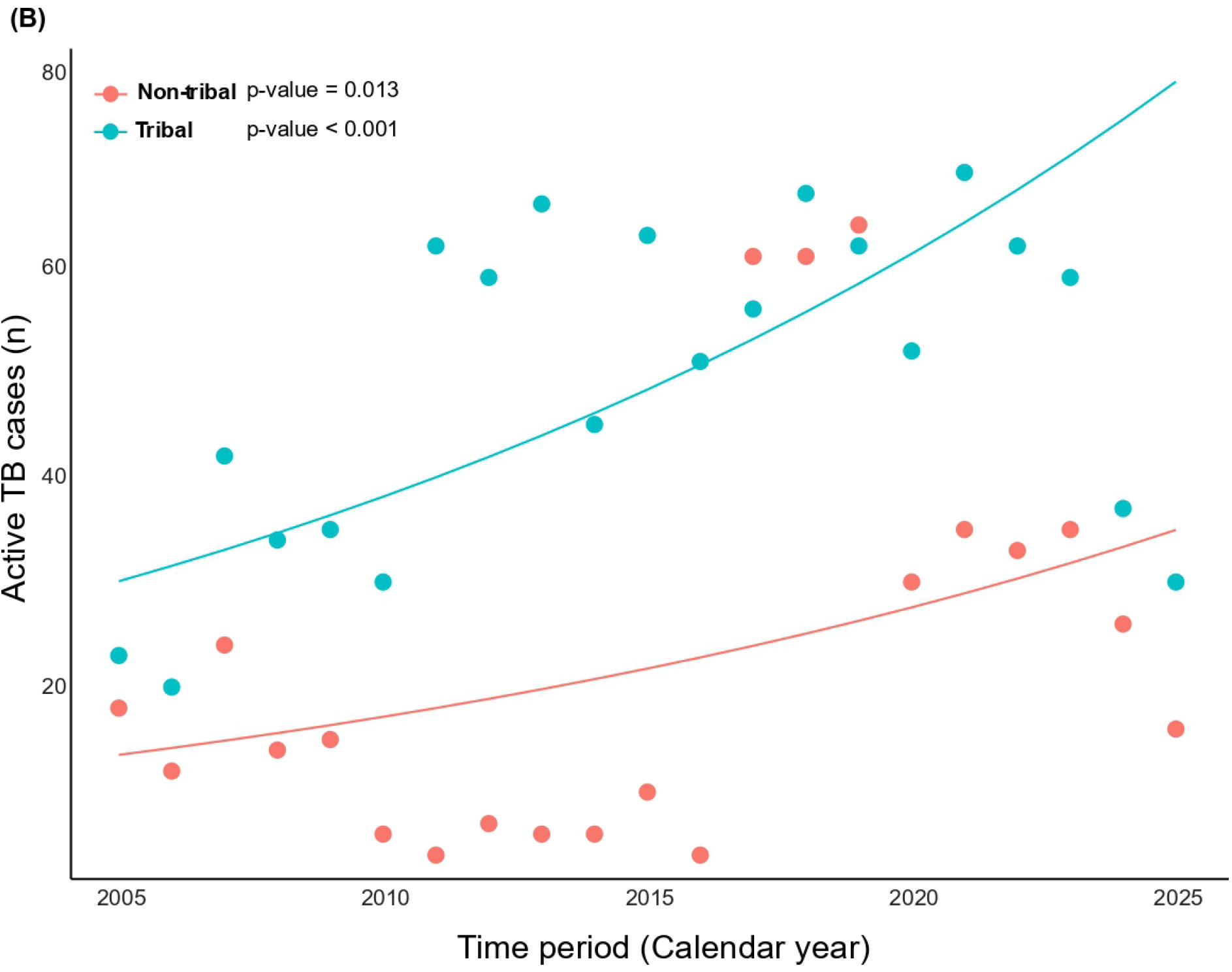
Poisson regression analysis of annual tuberculosis (TB) case counts among tribal and non-tribal populations. Points represent observed yearly TB cases, while solid lines represent fitted Poisson regression trends. Tribal populations consistently showed higher TB case counts than non-tribal populations throughout the study period. TB case trends differed significantly between tribal and non-Tribal populations over the study period (P < 0.001)

### Demographic and clinical characteristics

Of the 1,512 patients with TB included in the analysis, 1,021 (67.5%) were from tribal communities. Male patients constituted 58.2% of the tribal cohort with median age 33 years (IQR; 25-45), whereas the median age among tribal female patients was 25 years (IQR; 20-35) (**Table 1**). The majority of patients were newly diagnosed (86.2%), and pulmonary TB was the predominant clinical form (81.3%). Cough was the most frequently documented presenting symptom (93.8%). Smear microscopy results were available for 78.7% of patients, and GeneXpert MTB/RIF was performed in 36.2% of cases. Overall, 53% of patients were classified as cured and 27% as having completed treatment, while 20% experienced unfavorable treatment outcomes (**Table 2**).

**Table 1:** Socio-demographic and clinical characteristics of tribal and non-tribal tuberculosis patients. Data are presented as number (percentage) for categorical variables and median for age. The table summarizes patient demographics, nutritional status, clinical symptoms, radiological findings, microbiological results, disease characteristics, case type, and treatment outcomes according to population group.

| Parameter | Total (n = 1512) | Tribal (n = 1021) | Non-Tribal (n = 491) |
| --- | --- | --- | --- |
| Male (n (%)) | 888 (58.7%) | 594 (58.2%) | 294 (59.9%) |
| **Age (Years) (median)** |  |  |  |
| 0-15 | 122 (7) | 97 (7) | 25 (5) |
| 16-30 | 613 (24) | 464 (25) | 149 (24) |
| 31-45 | 449 (38) | 294 (38) | 155 (40) |
| 46-60 | 212 (52) | 115 (52) | 97 (53) |
| >60 yrs | 102 (65) | 46 (64.5) | 56 (68) |
| Not available | 14 (0.9%) | 5 (0.5%) | 9 (1.8%) |
| **BMI (kg/m2)** |  | **1021-710=311** | **491-376=115** |
| □<□18.5 | 358(84.0%) | 277 (89.0%) | 81 (70.4%) |
| □≥□18.5 and <□25 | 61 (14.3%) | 33 (10.6%) | 28 (24.3%) |
| □≥□25 | 7 (1.6%) | 1 (0.32%) | 6 (5.2%) |
| Not available | 1086 (71.8%) | 710 (69.5%) | 376 (76.6%) |
| **Symptoms** |  | **1021-717=304** | **491-330=161** |
| Cough | 434 (27.7%) | 285 (93.8%) | 149 (92.5%) |
| Fever | 51 (3.3%) | 29 (9.5%) | 22 (13.7%) |
| weight loss | 27 1.7%) | 18 (5.9%) | 9 (5.6%) |
| Night sweats | 5 (0.3%) | 3 (0.9%) | 2 (1.2%) |
| Not available | 1047 (66.9%) | 717 (70.2%) | 330 (67.2%) |
| **Chest-Xray** |  | **1021-895=126** | **491-431=60** |
| Positive | 186 (12.3%) | 126 (100%) | 60 (100%) |
| Not available | 1326 (87.7%) | 895 (87.7%) | 431 (42%) |
| **Smear microscopy** |  | **1021-429=592** | **491-286=205** |
| AFB3+ | 387 (25.6%) | 304 (51.4%) | 83 (40.5%) |
| AFB2+ | 131 (8.7%) | 92 (15.5%) | 39 (19%) |
| AFB1+ | 182 (12.0%) | 127 (21.5%) | 55 (26.8%) |
| Scanty | 97 (6.4%) | 69 (11.7%) | 28 (13.7%) |
| Negative | 310 (20.5%) | 212 (35.8%) | 98 (47.8%) |
| Not available | 405 (26.8%) | 217 (21.3%) | 188 (38.3%) |
| **CBNAAT** |  | **1021-754=267** | **491-353=138** |
| RIF sensitive | 378 (93.3%) | 249 (93.3%) | 129 (93.5%) |
| RIF resistant | 14 (3.5%) | 10 (3.7%) | 4 (2.9%) |
| RIF-resistant indeterminate | 13 (3.2%) | 8 (3%) | 5 (3.6%) |
| MTB not detected | 182 (31%) | 103 (27.8%) | 79 (36.4%) |
| Not available | 925 (61.2%) | 651 (63.8%) | 274 (55.8%) |
| **FLLPA** |  | **1021-836=185** | **491-415=76** |
| RIF-S, INH-S | 252 (96.5%) | 180 (97.3%) | 72 (94.7%) |
| RIF-S, INH-R | 7 (2.7%) | 4 (2.2%) | 3 (3.9%) |
| RIF-R, INH-R | 2 (0.8%) | 1 (0.5%) | 1 (1.3%) |
| MTB not detected | 13 (4.7%) | 12 (6%) | 1 (1.3%) |
| Not available | 1238 (81.9%) | 824 (80.7%) | 414 (84.3%) |
| **Case type** |  | **1021-4=1017** | **491-4=487** |
| New | 1301 (86.0%) | 877 (86.2%) | 424 (87%) |
| Relapse | 164 (10.9%) | 107 (10.5%) | 57 (11.7%) |
| Retreatment | 39 (2.6%) | 33 (3.2%) | 6 (1.2%) |
| Not available | 8 (0.5%) | 4 (0.4%) | 4 (0.8%) |
| **Treatment outcome** |  | **1021-36=985** | **491-105=386** |
| Cured | 710 (47.0%) | 523 (51.2%) | 187 (48.4%) |
| Treatment completed | 411 (27.2%) | 266 (27%) | 145 (37.6%) |
| Treatment failure | 15 (1%) | 13 (1.3%) | 2 (0.5%) |
| Default | 108 (7.1%) | 95 (9.6%) | 13 (3.4%) |
| Death | 122 (8.1%) | 86 (8.7%) | 36 (9.3%) |
| On treatment | 5 (0.3%) | 2 (0.2%) | 3 (0.8%) |
| Not available | 141 (9.3%) | 36 (3.5%) | 105 (21.4%) |

**Table 2:**
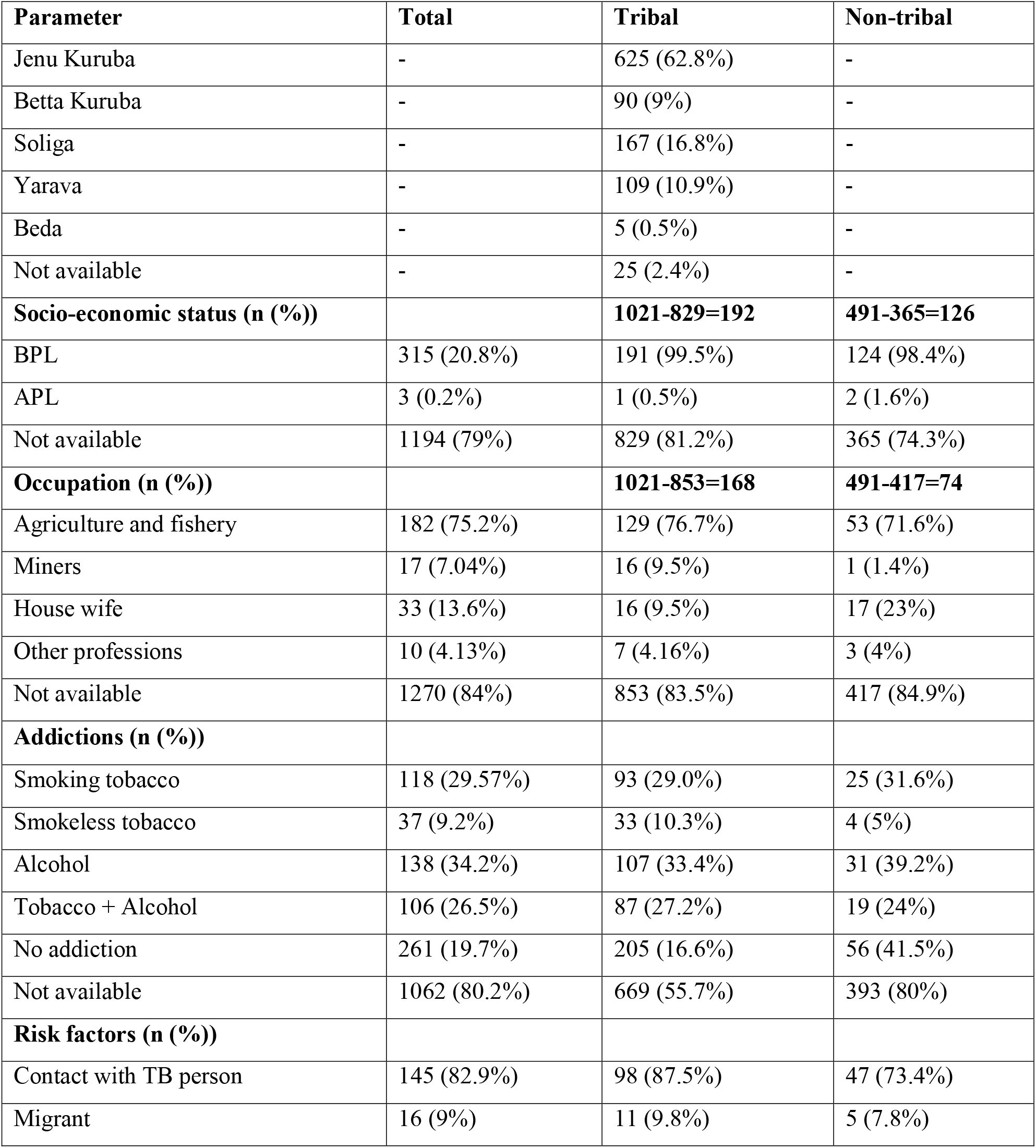
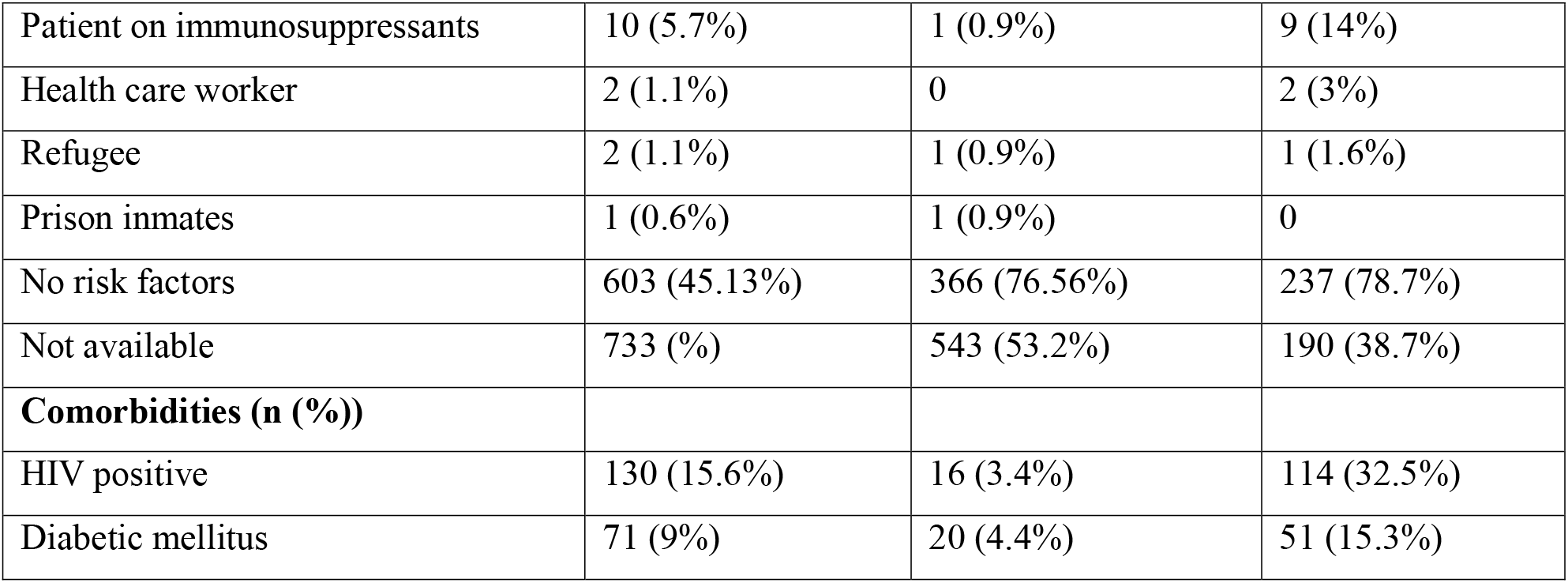
Sociodemographic characteristics, tribal subgroups, socioeconomic status, occupation, addiction history, risk factors, and comorbidities among tuberculosis patients. Data are presented as number (percentage). The table summarizes the distribution of tribal subgroups, socioeconomic status, occupation, behavioral risk factors, and comorbid conditions among tribal and non-tribal patients.

### Case reporting and data completeness

We observed that even before Nikshay implementation, TB notifications increased significantly over time in tribal communities (count ratio [CR] 1.064, 95% CI 1.032-1.097; p<0.001), an average annual rise of about 6.4% before 2018.

At Nikshay implementation, notifications rose immediately (CR 1.51, 95% CI 1.07-2.13; p=0.019), about 51% higher than expected from the pre-intervention trend. After this level change, the slope declined significantly (CR 0.889, 95% CI 0.836-0.945; p<0.001), an average annual decrease of about 11.1% after 2018. Notifications nonetheless remained above the pre-2018 baseline because of the initial increase. Sensitivity analyses excluding the intervention year (2018) and the COVID-19 year (2020) yielded similar estimates and conclusions (**Figure 2A**). DHARMa residual diagnostics indicated adequate model fit, with no significant deviation from the expected distribution (Kolmogorov-Smirnov p=0.657), no over- or underdispersion (p=0.984), and no outliers (p=1.00) (Figure 2B).

**Figure 2(A):**
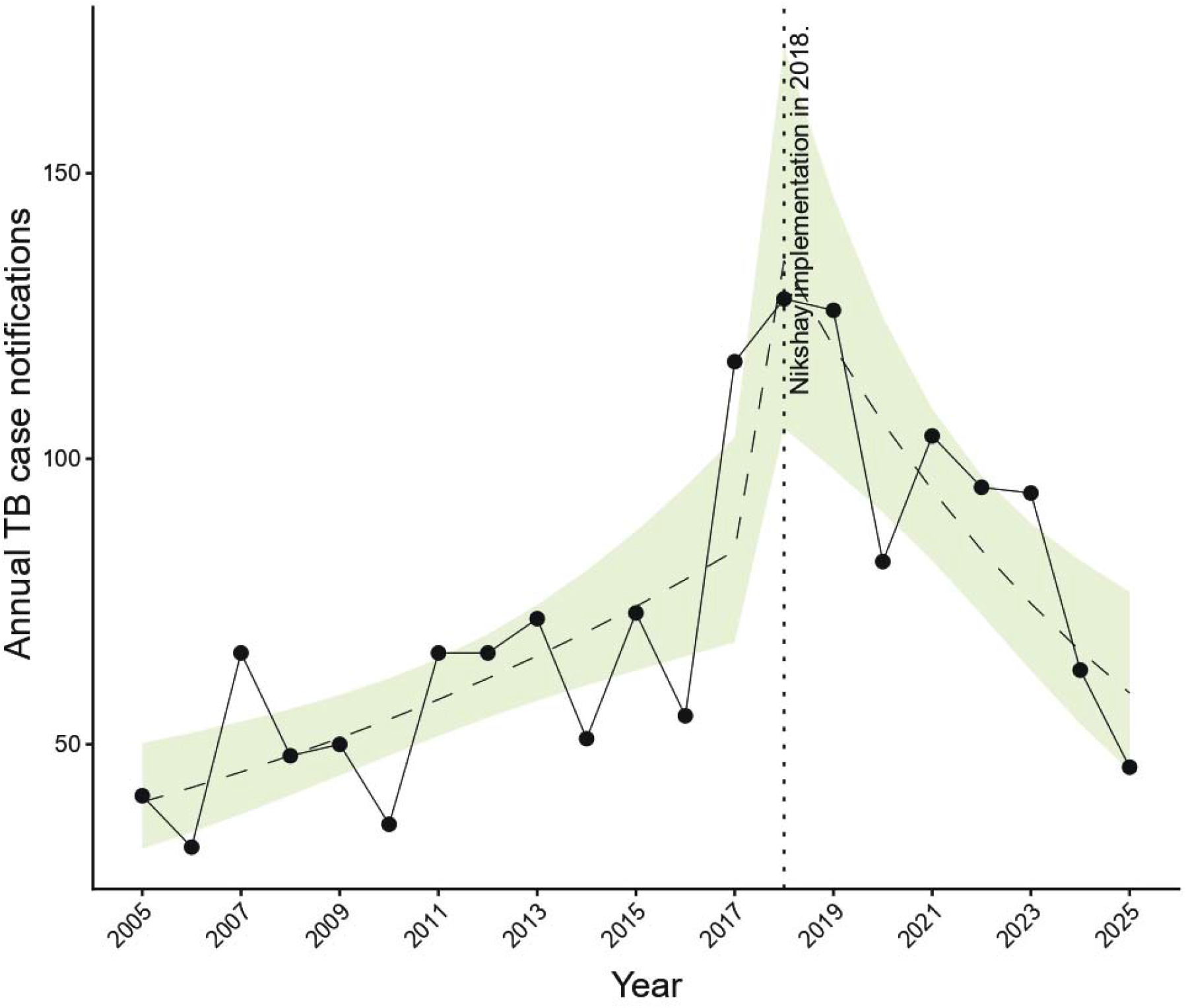
Annual trend in tuberculosis cases reported among tribal and non-tribal populations in the study area (2005–2025). Stacked bars represent the annual number of tuberculosis (TB) cases reported among tribal (green) and non-tribal (orange) populations. The dashed vertical line indicates the introduction of the Nikshay portal in 2018. Grey bars denote years influenced by active HIV screening (2017) and the COVID-19 pandemic (2020), which may have affected TB case detection and reporting.

**Figure 2(B):**
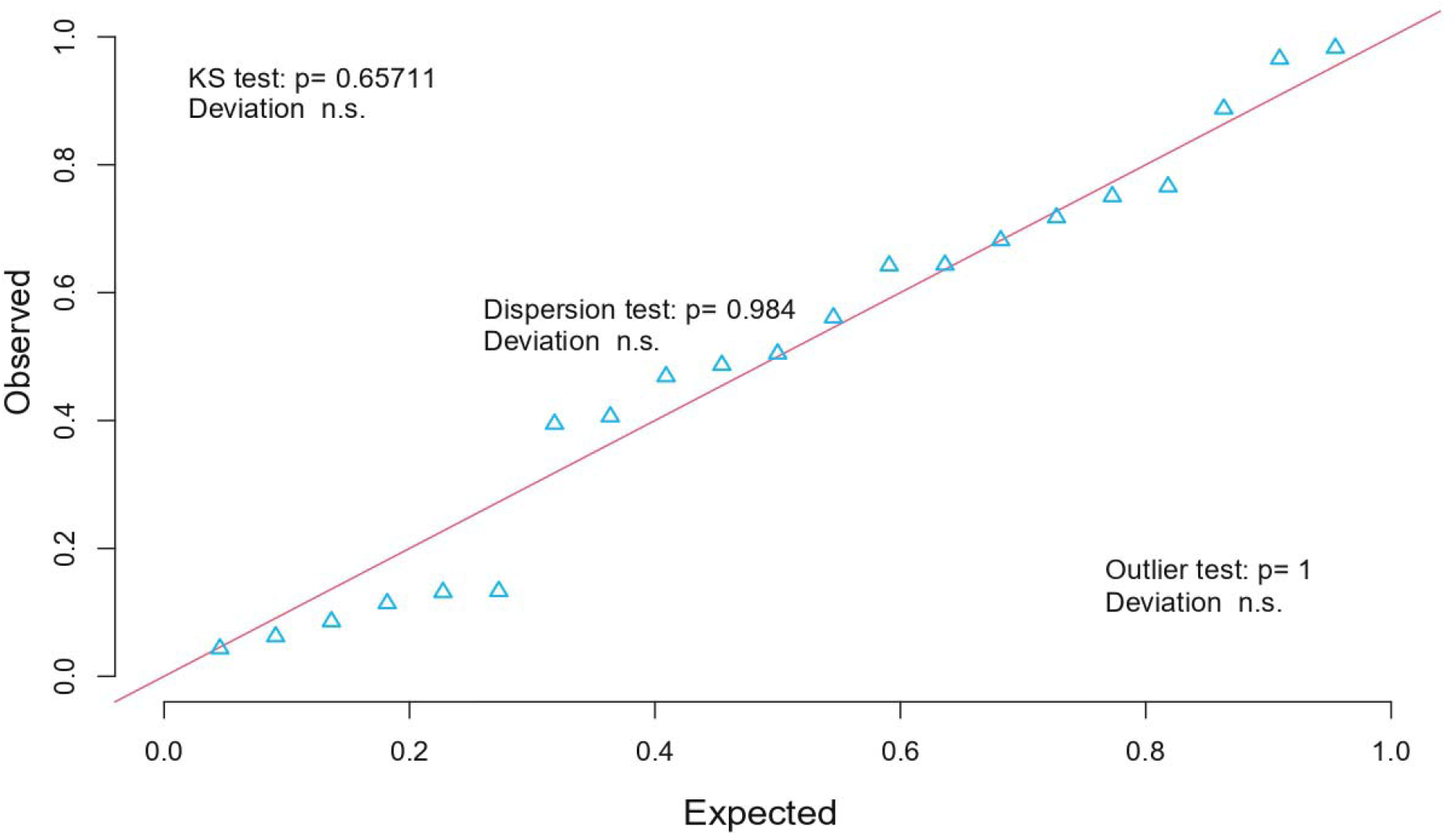
DHARMa model diagnostics based on simulated residuals indicated no evidence of overdispersion (dispersion test, *p* = 0.98) or a significant deviation from the expected uniform residual distribution (Kolmogorov–Smirnov test, *p* = 0.65711).

Data completeness improved markedly after Nikshay implementation. Body weight was recorded for 14.0% of patients in hospital registers versus 89.9% in Nikshay; HIV status for 11.5% versus 98.4%; and diabetes status for 9.5% versus 96.7%. Presenting symptoms were available for 0.5% of records before Nikshay and 62.5% after, while case type and treatment outcomes were captured consistently (>99%) in both periods. Occupation, addictions, household contact-tracing, socio-economic status, diagnostic tests, and other risk factors were routinely recorded only after Nikshay. Overall completeness was significantly higher in Nikshay (W=9900; p<0.001) (**Figure 3; Supplementary Table 1).**

**Figure 3:**
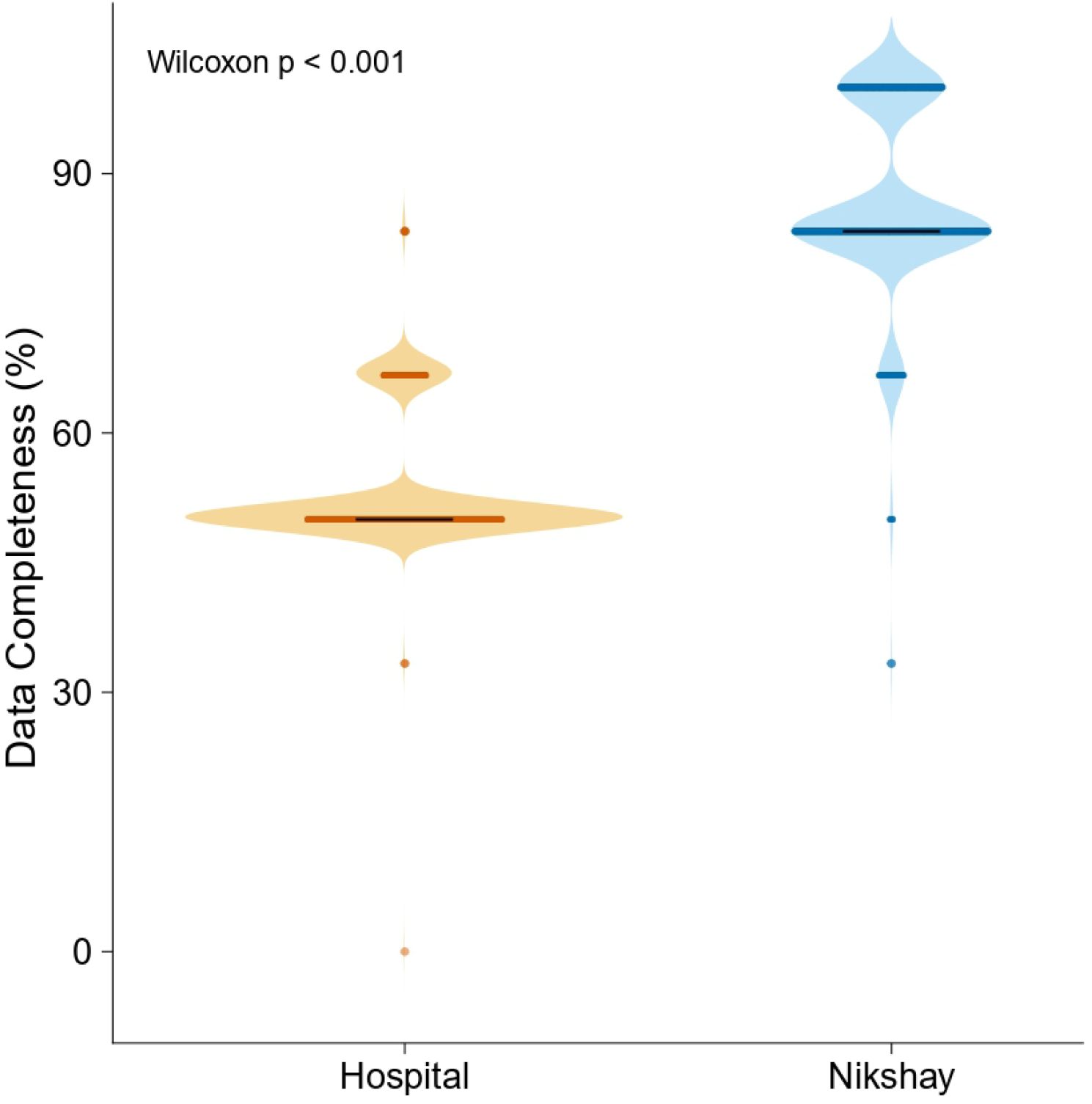
Distribution of variable completeness in hospital records and the Nikshay portal. Violin plots represent the distribution of completeness (%) across all study variables, whereas each point in the beeswarm plot corresponds to the completeness of an individual variable. The figure demonstrates the variability in data availability between the two data sources and identifies variables with high and low completeness.

### TB and malnutrition in tribes

Baseline BMI data were available for 426 of 1512 patients (28.2%), including 311 of 1021 tribal individuals (30.5%) and 115 of 491 non-tribal individuals (23.4%). The median baseline BMI was 15.3 kg/m² (IQR; 3.9). Malnutrition was highly prevalent, particularly among tribal patients. Overall, 360 of 426 (84.5%) were underweight (BMI <18.5 kg/m²), with a significantly higher proportion in tribal patients (89.4%) compared with non-tribal patients (71.3%) (p<0.001).

### Comorbidities

Comorbidity data were available for 819 patients (Tribal cases, n = 467 (57%) and non-tribal cases, n = 352 (43%)). HIV infection and diabetes mellitus (DM) were documented in 130 (15.9%) and 71 (8.7%) individuals, respectively. HIV co-infection was disproportionately concentrated in the non-tribal population, accounting for 114 of 130 cases (87.7%), with only 16 cases (12.3%) among tribal patients. A similar distribution was observed for DM, with 51 of 71 cases (71.8%) occurring in non-tribal individuals compared with 20 (28.2%) in tribal individuals.

### Addiction patterns, host risk factors, and their impact on TB treatment outcome

Treatment outcomes were available for 1,371 TB patients: 710 (51.8%) were cured, 411 (30.0%) completed treatment, and 5 (0.4%) remained on treatment at analysis. Overall, 108 (7.8%) were lost to follow-up (tribal 9.6%, non-tribal 3.4%), 15 (1.1%) had treatment failure (1.3% vs 0.5%), and 122 (8.9%) died (8.7% vs 9.3%).

Addiction data were available for 450 patients: 49 (10.9%) used tobacco only, 32 (7.1%) alcohol only, and 106 (23.6%) both, while 261 (58.4%) reported none. In the complete-case multivariable logistic regression (784 patients, 94 with poor outcomes), male sex (aOR 2.81, 95% CI 1.70-4.85; p<0.001), tribal status (aOR 1.85, 95% CI 1.10-3.19; p=0.023), and diabetes (aOR 2.10, 95% CI 1.05-4.03; p=0.029) were independently associated with poor outcome. HIV infection was not (aOR 1.38, 95% CI 0.65-2.78; p=0.383). No problematic multicollinearity was observed (all variance inflation factors <1.4), and model calibration was acceptable (**Figure 4**).

**Figure 4:**
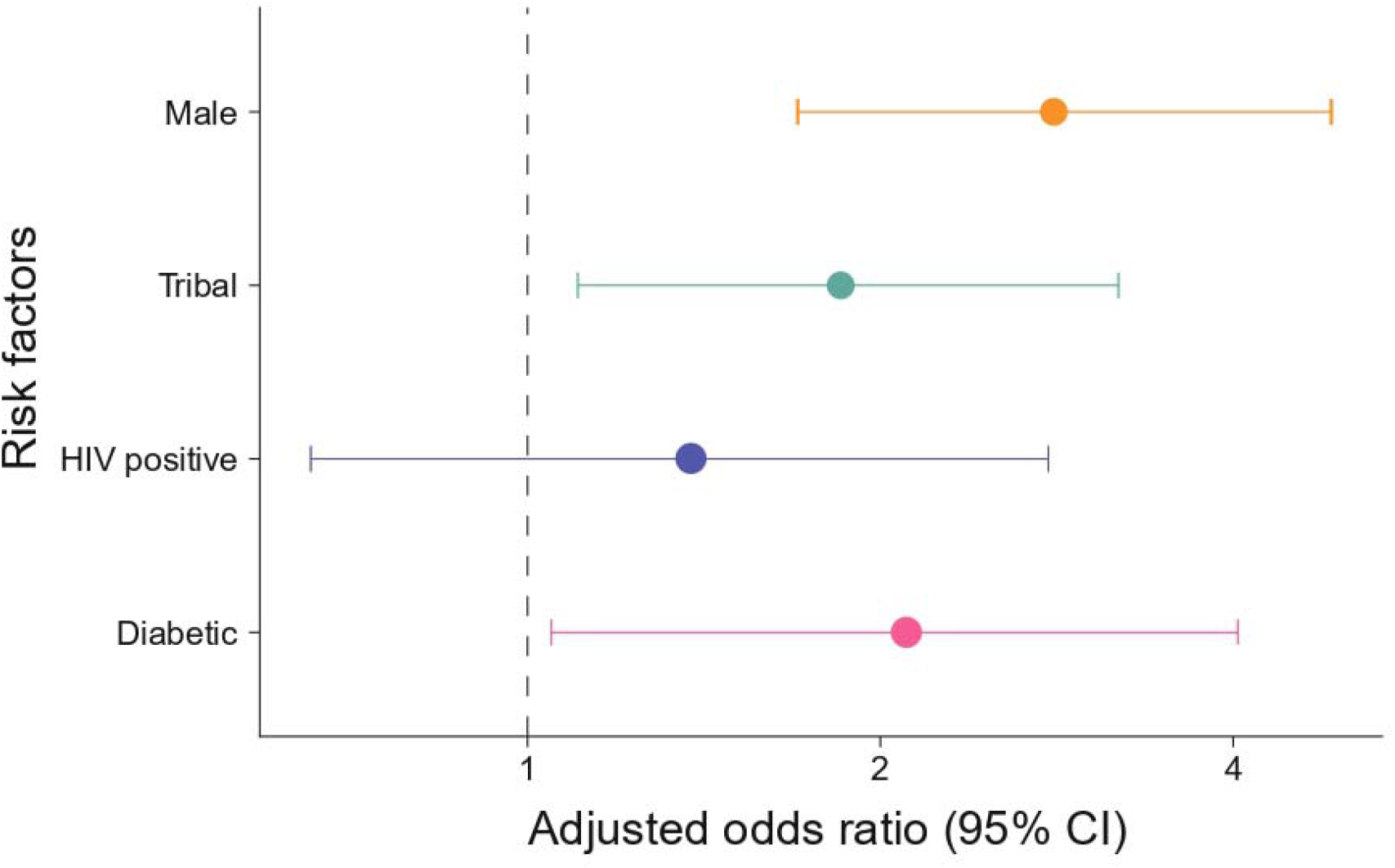
Multivariable logistic regression analysis of factors associated with poor tuberculosis treatment outcomes. The forest plot presents adjusted odds ratios (aORs) and 95% confidence intervals for each predictor included in the model. Points represent adjusted odds ratios, and horizontal lines indicate 95% confidence intervals. The vertical reference line (OR = 1) denotes no association between the predictor and poor treatment outcome.

### Household contact investigation and screening outcomes

After Nikshay implementation, 503 index TB cases yielded 1,535 household contacts (1,019 [66.4%] tribal; 516 [33.6%] non-tribal). Of these, 1,161 (75.6%) were screened; 80 (6.9%) were symptomatic and 17 (1.5%) were diagnosed with active TB (12 tribal, 5 non-tribal). Treatment was initiated for 11 tribal and all 5 non-tribal diagnosed contacts. The number needed to screen was 68.3. Screening coverage was significantly lower among tribal than non-tribal contacts (69.8% vs 87.2%; χ²=55.56, p<0.001), indicating gaps in contact investigation despite higher yield in tribal households.

## DISCUSSION

TB disproportionately affects Indigenous tribal communities. In this 21-year retrospective study from Karnataka in India, we analyzed active TB cases among tribal and neighbouring non-tribal populations attending a common referral hospital. TB incidence, poorly documented in these tribal groups, rose over time in both populations, more steeply among tribal communities. Introduction of the Nikshay digital reporting system markedly improved data completeness for key variables, including body weight, HIV and diabetes status, symptoms, substance use, occupation, and contact tracing, underscoring the value of standardized electronic reporting for TB surveillance in hard-to-reach tribal populations.

Comparable digital systems have strengthened infectious disease surveillance elsewhere. Electronic platforms for HIV and hepatitis C improved data handling and treatment monitoring^16^, while China’s CIDARS enabled automated outbreak detection and rapid response^17^. Korea’s integrated surveillance system similarly enhanced evidence based policymaking^18^. In our setting, Nikshay improved capture of critical variables such as nutritional status, drug resistance, and behavioural risks, enabling more granular epidemiological assessment.

High TB incidence among Indigenous populations is well documented, often exceeding national averages. For example, the Saharia tribe reported an annual incidence of 1504 per 100,000^19,20^, and national estimates place tribal pulmonary TB prevalence between 432 and 894 per 100,000^6^. Consistent with this evidence, we observed a faster rise in TB incidence among tribal communities, suggesting persistent vulnerability and potential for ongoing transmission.

Undernutrition remains a critical driver. Chronic energy deficiency exceeds 40-45% in some tribes^14^, and low BMI is widely reported^21,22^. In our cohort, 89.4% of tribal patients had low BMI. Documentation improved markedly after Nikshay implementation, with BMI recording increasing from 14% to 89.9%. Such improvements are essential, as nutritional status strongly influences TB susceptibility and outcomes. Behavioral and clinical factors further shaped outcomes. Tobacco and alcohol use were common, and smoking independently predicted poor treatment outcomes, consistent with meta analysis^23^. HIV co infection and diabetes also increased the risk of unfavourable outcomes, underscoring the need for integrated TB-HIV and TB-diabetes care. Social determinants including poverty, undernutrition, and limited healthcare access remain central^17,18^.

Tribal origin was associated with poorer outcomes, and lower household contact screening coverage indicates ongoing barriers to care, echoing known diagnostic gaps^6,24^. Although treatment success exceeded 74%, lost to follow-up, failure, and mortality persist, likely reflecting delayed diagnosis and structural inequities. The low number needed to screen among tribal contacts highlights contact investigation as a high yield strategy, though culturally appropriate implementation is essential^22,24^.

This study has limitations. Its retrospective design and reliance on routine data introduce potential information bias, particularly before Nikshay implementation. Being a single centre study, findings may not generalize to all tribal populations. Finally, causal relationships cannot be established, and prospective studies are needed to validate these findings.

## CONCLUSIONS

Nikshay implementation improved the completeness and detail of TB surveillance among Indigenous tribal communities in Karnataka, enabling better assessment of disease burden, risk factors, and treatment outcomes. Strengthening gender-sensitive, tribe-specific services, integrating TB-HIV and TB-diabetes care, expanding rapid molecular diagnostics, and incorporating tobacco and alcohol cessation into TB programs are essential. Future prospective studies using Nikshay’s digital platform, with detailed data on nutrition, adherence, and household conditions, are needed to guide targeted interventions and reduce TB morbidity and mortality in tribes.

## Data Availability

All data produced in the present study are available upon reasonable request to the authors

## Author Contributions

RV, KGS and AP conceptualized and designed the study. MR, ASB, and AR contributed equally to data collection and curation. MR, RV, ASB, KGS, AR, SV and PP performed the data analysis. PP and SV contributed to data analysis, interpretation of the findings, and critically reviewed the manuscript. RV, MR and ASB drafted the manuscript. All authors reviewed the manuscript, provided intellectual input, approved the final version.

## Acknowledgements

The authors sincerely thank the staff of the Vivekananda Memorial Hospital and the tuberculosis program for their support in data collection and access to the retrospective records.

## Data availability

The data that support the findings of this study are available on request from the corresponding author.

