## Supplementary table 1 for "IMPACT OF CENTRALIZED NIKSHAY ON TB NOTIFICATION AND DATA COMPLETENESS IN INDIGENOUS TRIBES: A 21-YEAR STUDY"

| **Variable** | **Hospital records n/N (%) complete** | **Nikshay n/N (%) complete** | **Difference (%)** | **P-value** |
| --- | --- | --- | --- | --- |
| Age | 350/400 (87.5) | 760/770 (98.7) | +11.2 | <0.001 |
| Sex | 378/400 (94.6) | 768/770 (99.8) | +5.2 | <0.001 |
| Occupation | 232/400 (57.9) | 703/770 (91.3) | +33.4 | <0.001 |
| HIV status | 266/400 (66.4) | 738/770 (95.9) | +29.5 | <0.001 |

Supplementary Table 1: **A variable-by-variable completeness table with denominators**
